# Challenges in Plasmodium diagnostics in countries nearing malaria elimination: a cross-sectional survey among treatment-seeking patients in health facilities in malaria endemic provinces of Cambodia with contrasted transmission intensity

**DOI:** 10.64898/2026.03.03.26347480

**Authors:** Nimol Khim, Agnes Orban, Sopheany Thin, Sitha Sin, Suzanne Guepin, Lionel Brice Feufack-Donfack, Virak Eng, Malen Ea, Sophy Chy, Chandavin Seng, Rotha Eam, Chanra Khean, Chanvong Kul, Nimol Kloeung, Sopheakvatey Ke, Claude Flamand, Michael White, Dysoley Lek, Jean Popovici

## Abstract

**Background:** Cambodia has made great progress in reducing malaria transmission and is targeting elimination. While this progress is particularly marked for *Plasmodium falciparum*, the situation is different for *Plasmodium vivax*. It is generally assumed that symptomatic patients are effectively diagnosed using rapid diagnostic tests (RDTs), regardless of transmission intensity.

**Methods:** In 2023 we conducted a cross-sectional survey among 986 treatment-seeking patients in 6 provinces of Cambodia with varying reported malaria cases. Malaria RDT (Pf/Pv), microscopy and qPCR diagnostics of *Plasmodium* infections and species determination were performed.

**Results:** Using qPCR, *Plasmodium* infections were diagnosed in 156 patients (15.8%, 95% CI: 13.7-18.2%) from all 6 provinces. Positivity rate was markedly different between health centers (HCs) and ranged between 57.2% and 0.5%. Parasitemia of infected patients was different between HCs and was lower in HCs with the lowest positivity rate compared to those with higher rates. The majority of *Plasmodium* infections (75%) were caused by *P. vivax*, however all human malaria species were identified as well as the simian parasite *P. knowlesi*. Overall sensitivity of RDTs to detect *Plasmodium* infections was 39.7% (95% CI: 28.9-51.6%) and specificity was 100% (95% CI: 99.5-100%). The proportion of RDT true positives was significantly different between HCs, and a tendency for higher false negative rates in low transmission areas compared to higher ones was observed.

**Conclusion:** While our results confirm that *P. falciparum* parasites are nearly eliminated from Cambodia, we show that current practice for diagnosis of *Plasmodium* infections among febrile patients is challenged, especially in very low transmission settings.

## BACKGROUND

Cambodia has made huge progress towards malaria elimination in the past 5 years with *Plasmodium falciparum* now almost eliminated from the country. In 2022, only 396 cases of *P. falciparum* were reported, a massive decrease compared to the 10000 cases reported in 2018 (1).

This reduction in malaria incidence is the result of intensive measures conducted by the Cambodian Malaria Control Program, such as chemoprophylaxis for at-risk individuals, reactive case detection upon diagnosis of any *P. falciparum* case and implementation of a network of community health workers trained for malaria diagnosis by rapid diagnostic tests (RDTs) and prompt and effective treatment for infected individuals (2–6). These efforts were initially aimed at eliminating *P. falciparum* defined as the priority. Although these interventions also reduced the incidence of the co-sympatric *P. vivax*, the effect was less pronounced, with 30680 cases in 2018 and nearly 4000 cases still reported in 2022 (1). This difference in response to control efforts relates to the fact that *P. vivax*, unlike *P. falciparum*, forms a dormant liver stage (hypnozoites) capable of causing multiple recurring blood stage infections (relapses) following a single mosquito inoculation thereby complicating vivax malaria elimination efforts (7). There is currently no diagnostic test for hypnozoite carriage. In addition, we, and others, have shown that asymptomatic parasite carriers represent the vast majority of *P. vivax* infections within endemic areas (8,9). Another factor that could contribute to the resilience of *P. vivax* to elimination efforts is a sub-optimal performance of RDTs used in most health centers (HCs) and by all community health workers as symptomatic *P. vivax* infections tend to present lower parasitemias compared to symptomatic *P. falciparum* (7).

In addition to the challenges of *P. vivax* elimination, Cambodia faces the issue of also being endemic for all other human malaria species (*Plasmodium ovale* and *Plasmodium malariae*) as well as several reports of zoonotic malaria caused by *Plasmodium knowlesi* (1,10,11). Performance of current diagnostic tools used in the country for these species is poorly described and often reported as insufficient (12–14). In order to obtain WHO malaria-free certification, countries need to demonstrate absence of malaria infections in its population, regardless of the *Plasmodium* species. Besides, plans for preventing re-establishment of malaria once a region is malaria-free need to be implemented and these rely on appropriate diagnosis of infections.

Although a substantial body of studies has assessed the diagnostic performance of microscopy and RDTs, the vast majority of these were carried out among patients infected by (i) *P. falciparum,* (ii) in high endemicity settings, (iii) single region and/or (iv) in asymptomatic carriers enrolled in community-based surveys (15–19). Here we report a cross-sectional survey conducted in six provinces of Cambodia where *Plasmodium* infections were systematically investigated using molecular diagnostics in symptomatic individuals seeking treatment in participating health centers.

## METHODS

### Study sites and population

This study was conducted in eight HCs distributed across six Cambodian provinces with significant malaria transmission (incidence in 2021-2022 from 0.27 to 1.62 cases per 1000 person per year): Kampong Speu (Aoral HC and Trapaing Cho HC), Pursat (Kravanh HC), Kratie (Kampong Cham HC) and Stung Treng (Siem Pang HC) or where malaria was historically endemic but where very few cases were reported in the past two years (incidence in 2021-2022 from 0.02 to 0.07 cases per 1000 person per year): Pailin (Kra Chab HC) and Battambang (Tasanh HC and Boeung Run HC) (Figure 1) (20).

**Figure 1.**
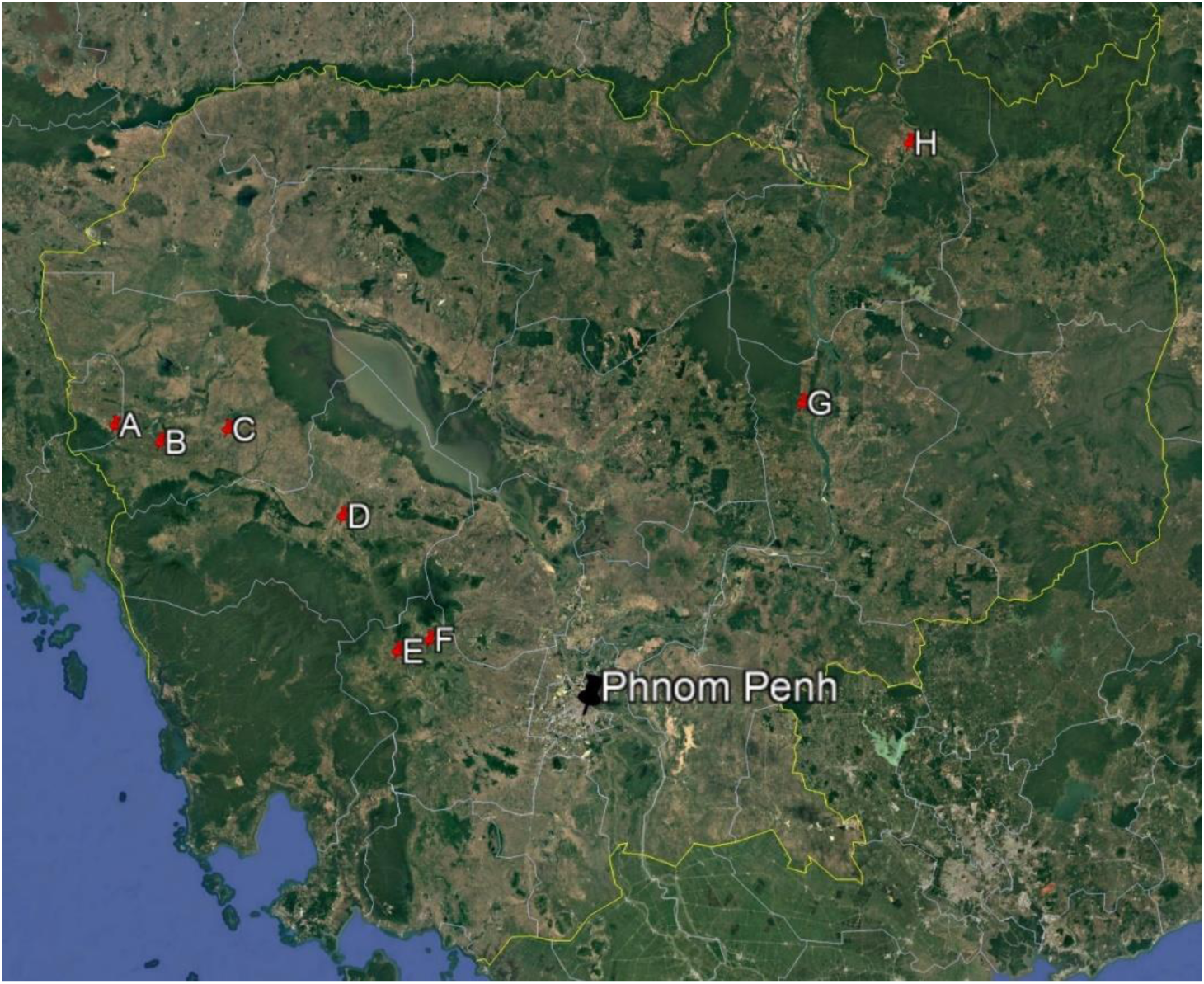
Location of HCs participating in the study. Eight HCs were selected across 6 provinces of Cambodia: Kra Chab HC (A) in Pailin, Tasanh HC (B) and Boeung Run HC (C) in Battambang, Kravanh HC (D) in Pursat, Aoral HC (E) and Trapaing Cho HC (F) in Kampong Speu, Kampong Cham HC (G) in Kratie and Siem Pang HC (H) in Stung Treng.

Between June 2023 and December 2023, individuals presenting to participating HCs, aged more than 5 years old with malaria-like symptoms (mainly febrile or with history of fever in the past 48h) were offered to participate. Among those, a number of participants (n=88) attended HC following a positive malaria RDT performed by Village Malaria Workers (VMW). Indeed, in Cambodia, G6PD testing and primaquine administration is performed only in HCs, therefore individuals seeking treatment at the community level and diagnosed by VMW with *P. vivax* infection are then referred to nearby HC (hereafter: pre-diagnosed). Upon consent of participants or their guardians, a capillary blood sample was collected by fingerprick into an EDTA microtainer tube, a thick and thin smear microscopy slide was prepared and a malaria RDT (Malaria Ag Pf/Pv Abbott) was performed by the HC staff. RDT storage, handling and testing was performed according to the national guidelines. RDT results were recorded and whole blood samples were stored at 4°C until being sent to Institut Pasteur du Cambodge in Phnom Penh within 72 h of collection. Sex and age of participants were also recorded. Patients with a positive malaria RDT result received standard care from the HC staff following Cambodian treatment guidelines.

### Laboratory procedures

DNA was extracted from 50 μl of blood for all samples using QIAamp DNA Mini Kit (QIAGEN) following the manufacturer’s instructions. A pan-*Plasmodium* qPCR targeting conserved regions of all *Plasmodium* cytB mitochondrial gene was performed to screen for any malaria infection (21). For *Plasmodium* positive samples, species-specific qPCRs were done to identify *P. falciparum*, *P. vivax*, *P. ovale* and *P. malariae* infections. *P. vivax* positive samples were then screened for *P. knowlesi* infections by species-specific qPCR. In case species could not be identified, a PCR targeting cytB and 18S genes were performed and amplicons were Sanger sequenced (Macrogen) (22–24). Sequences obtained were then used to identify the *Plasmodium* species using BLAST alignment tool (NCBI). All primers are listed in Supplementary Table 1.

All microscopy slides were stained using Giemsa staining (2%, 45 min) upon reception in the laboratory. Slides from all qPCR positive samples were read by counting 500 white blood cells (WBC) on the thick film and parasitemia was calculated assuming 8000 WBC per μl of blood.

### Outcomes

The primary outcome was to determine the positivity rates of *Plasmodium* infections among treatment-seeking patients using qPCR diagnostics. Secondary outcomes included the evaluation of risk factors associated to *Plasmodium* infections and the assessment of performance of RDTs and microscopy to diagnose malaria.

### Sample size

In 2022, across all Cambodia, malaria positivity rates in health facilities were nearly 1% (20). As these were diagnosed using RDTs, we estimated that expected positivity rates using a more sensitive diagnostic tool would be up to 5%. With a 2% margin of error and a Z-score of 1.96, the required sample size was 456. We applied a design effect factor of 2 to take into account variability across HCs leading to a final sample size of 912 rounded up to 1000.

### Statistical analyses

Differences in proportions was assessed by χ^2^ or Fisher’s tests as appropriate. Differences in medians between multiple categories was assessed by Kruskal-Wallis test. Correlations between quantitative variables was assessed by Spearman rank test. Differences between two categories in median of non-normally distributed variables was assessed by Mann-Whitney test.

Positivity rates of *Plasmodium* infections was calculated overall and stratified by key demographic variables including sex, age category and province. Pearson’s chi-squared test was used to assess associations between categorical variables. To identify factors independently associated with *Plasmodium* infection, we fitted multivariable Poisson regression models with robust variance estimation to directly estimate crude and adjusted infection risk ratios (IRRs) and 95% confidence intervals (95% CI). The global significance of each categorical variable included in the model was assessed using a Wald test for joint significance (testparm). Statistical significance was defined as a two-sided p-value <0.05 for the Pearson and Wald test or a 95% CI for IRRs. Data were analyzed using R software, version 4.5.0 (Ref R Core Team. R: A language and environment for statistical computing. Version 4.5.0. Vienna, Austria: R Foundation for Statistical Computing, 2025) and GraphPad Prism version 10.5.0.

### Ethics statement

This study was approved by the National Ethics Committee for Health Research of the Ministry of Health of Cambodia (#148 NECHR May 2^nd^ 2023).

## RESULTS

Between June and December 2023, 987 patients seeking treatment at participating HCs were enrolled in this study. Overall, the majority of patients attending HCs were male (651/987, 66%) compared to female patients (336/987, 34%) and this proportion varied significantly between the HCs (χ^2^, p<0.0001) with as low as 33% of males in Tasanh HC and up to 84% in Siem Pang (Table 1). Median age of patients was 30 years old overall (IQR: 22-39) and was significantly different between HCs, ranging from 25 years old (IQR: 18-37) in Kampong Cham to 37 years old (IQR: 22-46) in Tasanh (Kruskal-Wallis, p<0.0001). Age distribution was also different (χ^2^, p<0.0001) between all HCs with a proportion of patients aged less than 15 years old ranging from 0% in Aoral to 41% in Boeung Run (Table 1 and Supplementary Figure 1.).

**Table 1.**
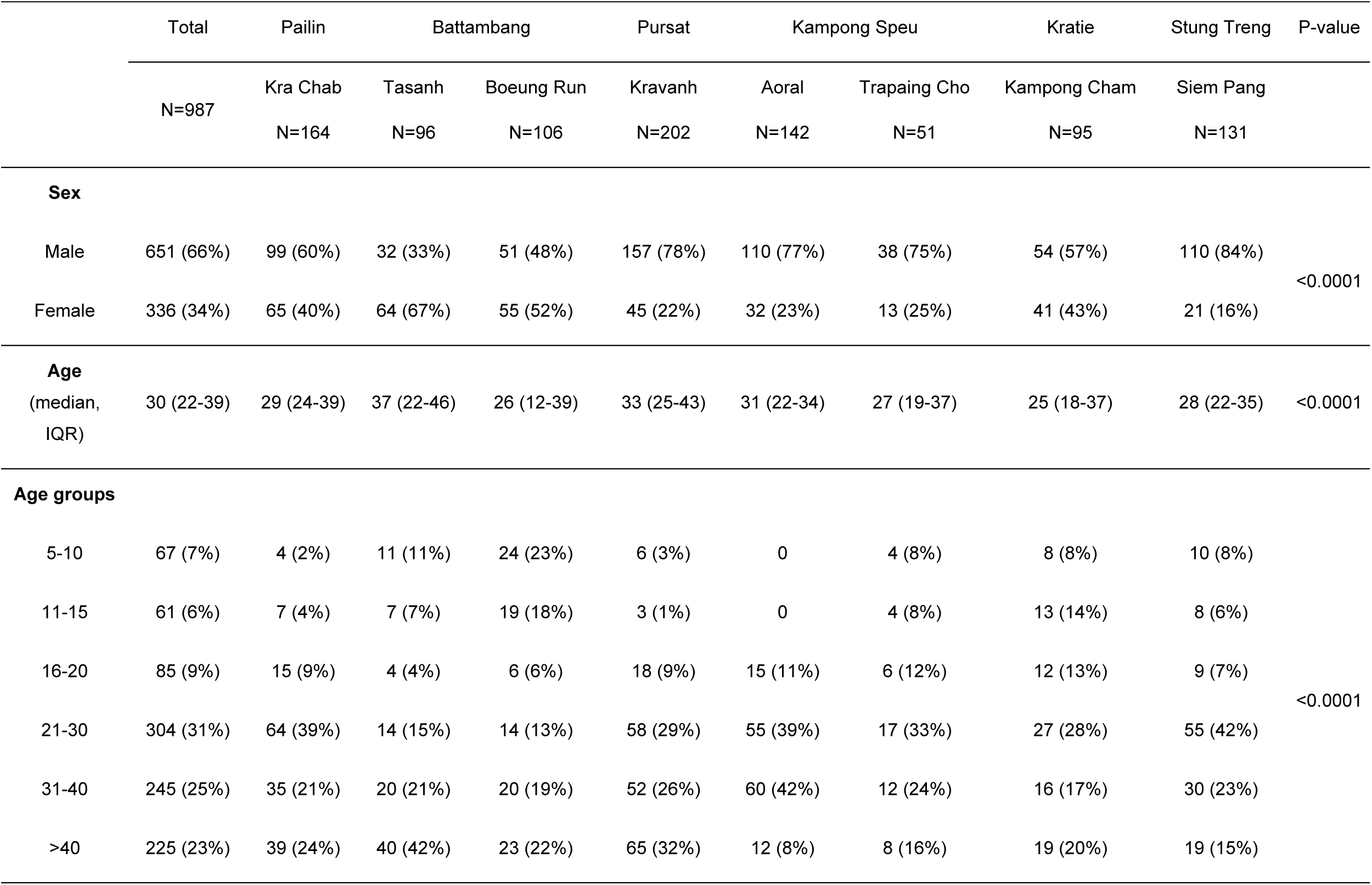
Baseline characteristics of patients recruited in the cross-sectional survey. Differences in proportions were analyzed using Chi-square tests, and the corresponding P-values are shown.

DNA extraction and qPCR diagnostics of *Plasmodium* infections could be performed on all but one of the 987 capillary blood samples collected from patients. Overall, *Plasmodium* infections were diagnosed by qPCR in 156 patients (15.8%, 156/986, 95% CI: 13.7-18.2%). The vast majority of infected patients were male (137/156, 88%) leading to an overall positivity rate of 21.0% (137/651, 95% CI: 18.1-24.3%) for males, significantly higher than 5.7% (19/335, 95% CI: 3.7-8.7%) for females (χ^2^, p<0.0001) (Table 2). There was also a significant difference in *Plasmodium* positivity rates according to the age of patients (χ^2^, p=0.0004). The majority of infected patients were aged 21 to 30 years old (69/156, 44%, 95% CI: 34.4-54.5%) and had the highest rates of *Plasmodium* infections (69/304, 22.7%, 95% CI: 18.3-27.7%). The lowest infection rate was in patients aged 5 to 10 years old (3/67, 4.5%, 95% CI: 1.2-12.4%). Considering only patients not pre-diagnosed by VMWs, infection rates in males was 9.5% (54/568, 95% CI: 7.4-12.2%), significantly higher than in females (14/330, 4.2%, 95% CI: 2.5-7.0%, χ^2^, p=0.0040). Due to an overall low number of infected patients in some age groups, there was no more significant difference in positivity rates according to the age of the patients not pre-diagnosed (χ^2^, p=0.3682). However, infection rate was significantly lower in children aged less than 15 (3/116, 2.6%, 95% CI: 0.7-7.3%) compared to patients aged more than 15 (65/782, 8.3%, 95% CI: 6.6-10.5%, Fisher, p=0.0243).

**Table 2.**
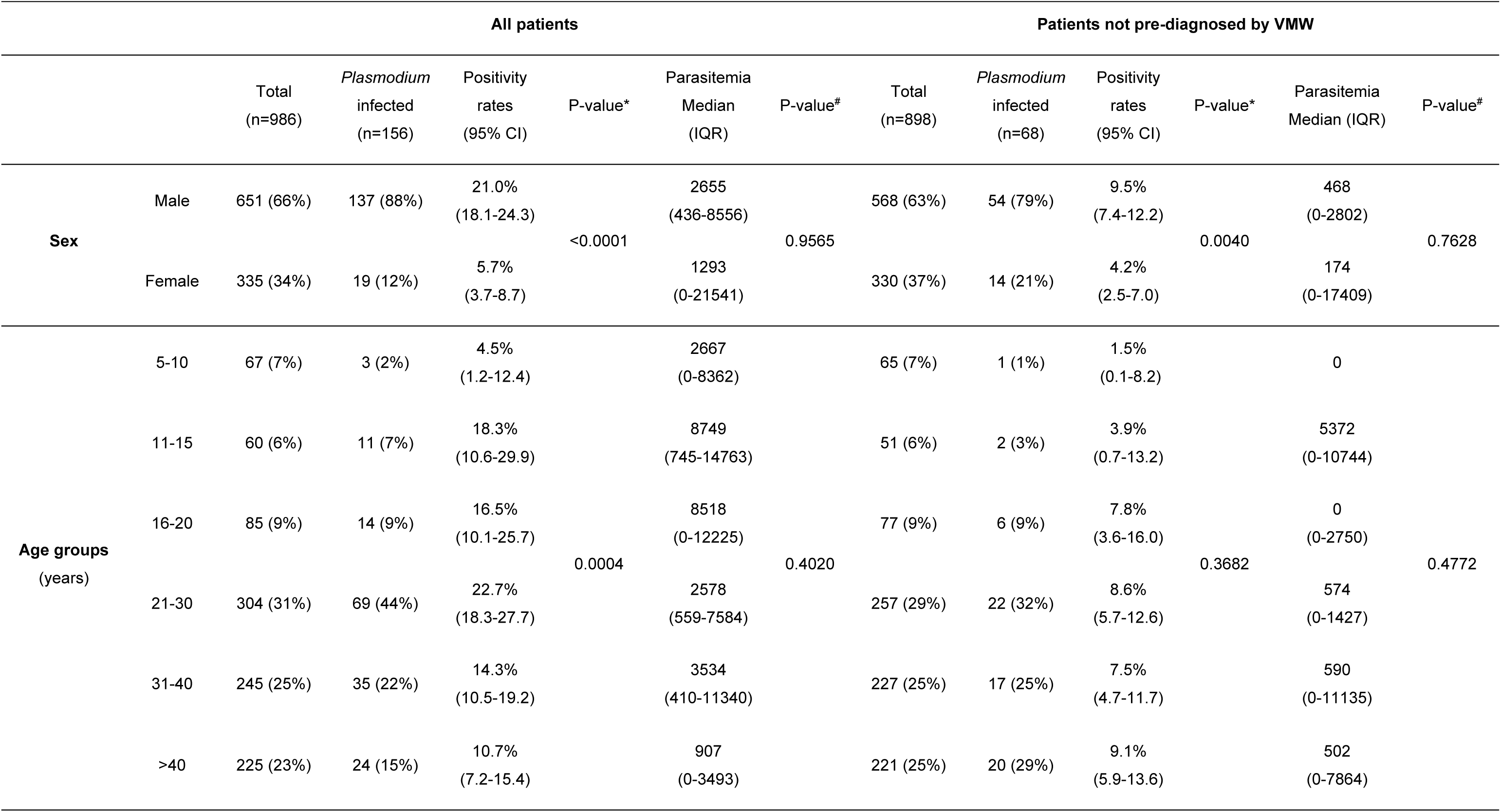
*Plasmodium* infections detected by RT-PCR among treatment-seeking patients. Differences in proportions were analyzed using Chi-square tests, and the corresponding P-values are shown. *: Comparison of positivity rates. ^#^: Comparison of parasitemia medians.

qPCR positive samples were found in all provinces although qPCR positivity rates (hereafter referred to as positivity rates) varied significantly between HCs (χ^2^, p<0.0001) (Table 3, Figure 2). The highest positivity rate was observed in Siem Pang HC in Stung Treng, where 57.2% (75/131, 95% CI: 48.7-65.4%) of patients tested positive. Conversely, Battambang exhibited the lowest rate of 0.5% (1/201, 95% CI: 0.0-2.8%), with Boeung Run HC reporting only a single case throughout the study period while no case was reported in Tasanh HC. In Kampong Speu, the overall positivity rate was 11.9% (23/193, 95% CI: 8.1-17.2%) with highly contrasted rates between the two HCs: 2.8% (4/142, 95% CI: 1.1-7.0%) in Aoral and 37.3% (19/51, 95% CI: 25.3-51.0%) in Trapaing Cho. Positivity rate in Pailin was 2.4% (4/164, 95% CI: 1.0–6.1%), 16.3% (33/202, 95% CI: 11.9–22.1%) in Pursat, and 21.1% (20/95, 95% CI: 14.1–30.3%) in Kratie.

**Figure 2.**
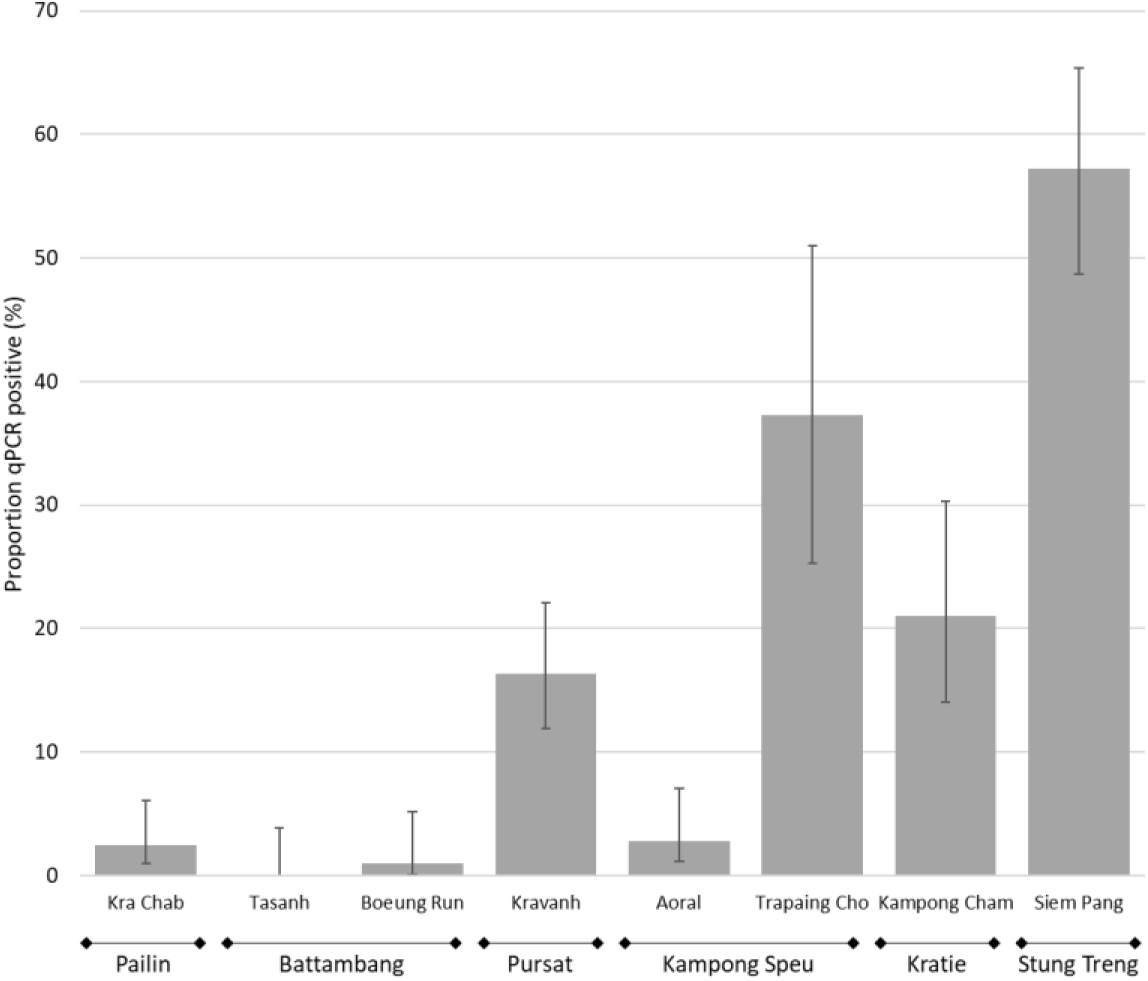
Positivity rates of *Plasmodium* infections detected by qPCR among all treatment-seeking individuals at the eight participating HC. Error bars represent 95% CI.

**Table 3.**
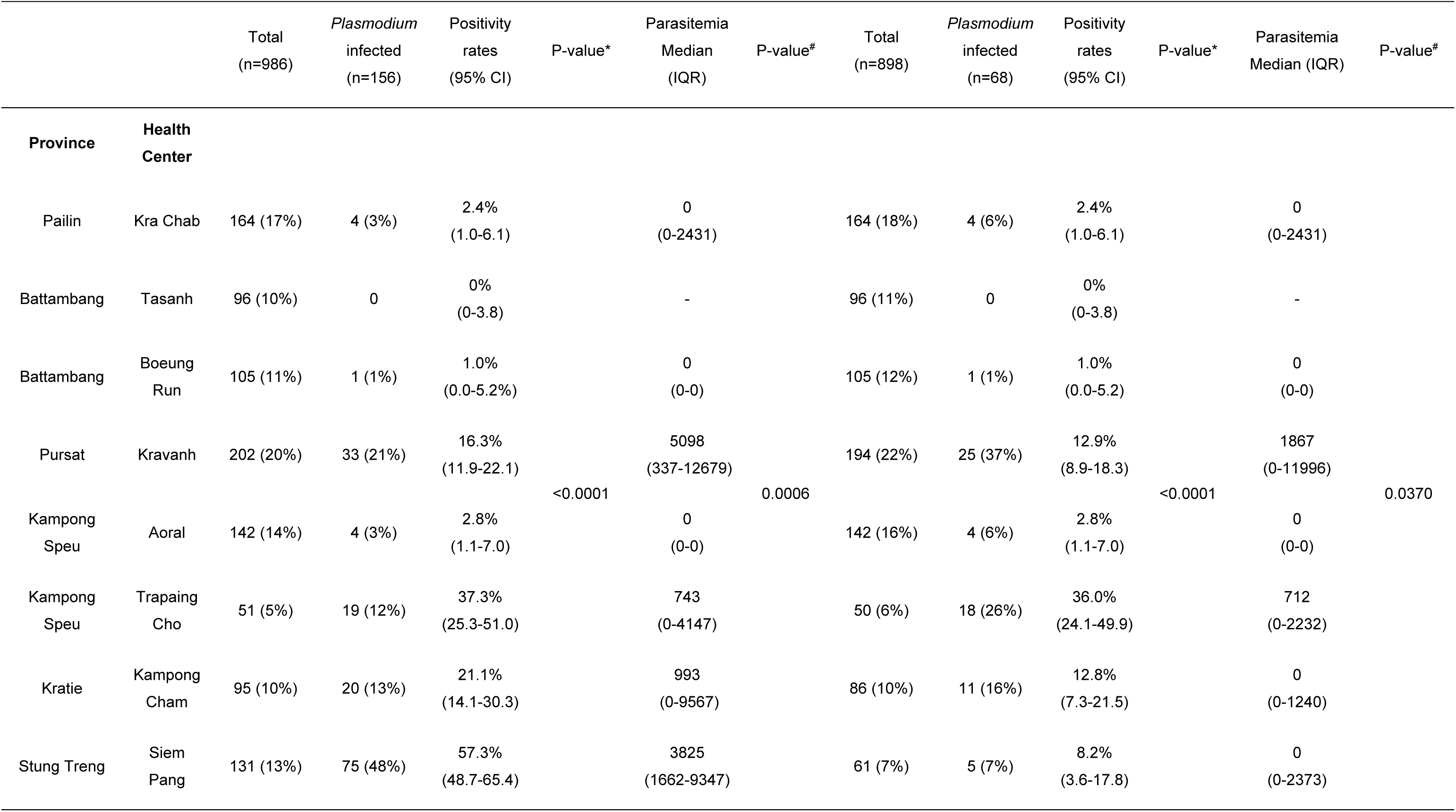
*Plasmodium* infections detected by RT-PCR among treatment-seeking patients. Differences in proportions were analyzed using Chi-square tests, and the corresponding P-values are shown. *: Comparison of positivity rates. ^#^: Comparison of parasitemia medians.

Overall positivity rate of *Plasmodium* infections among patients not pre-diagnosed by a positive RDT performed at the community level was 7.6% (68/898, 95% CI: 6.0-9.5%). As the majority of patients pre-diagnosed were from Stung Treng province (70/88), when considering only patients not pre-diagnosed, positivity rates remained identical or slightly lower for all HC except for Siem Pang HC in Stung Treng where it decreased to 8.2% (5/61, 95% CI: 3.6-17.8%) (Table 3).

Multivariable analysis confirmed that male sex was strongly associated with increased risk of *Plasmodium* infection (adjusted IRR = 2.14, 95% CI: 1.35-3.37%) (Table 4). Although crude infection rate was higher among participants aged 21-30 years old compared to those aged ≤15 years, age was not independently associated with infection in the adjusted model (p=0.23). Geographic location remained a strong predictor of infection. Compared to Kampong Speu, patients from Stung Treng had a markedly higher risk of infection (adjusted IRR = 4.63, 95% CI: 3.04-7.04%), followed by Kratie (adjusted IRR = 2.09, 95% CI: 1.20-3.64%), and Pursat (adjusted IRR 1.41, 95% CI: 0.84-2.35%). In contrast, patients from Battambang and Pailin had significantly lower risks (adjusted IRR = 0.06 and 0.23, respectively).

**Table 4.**
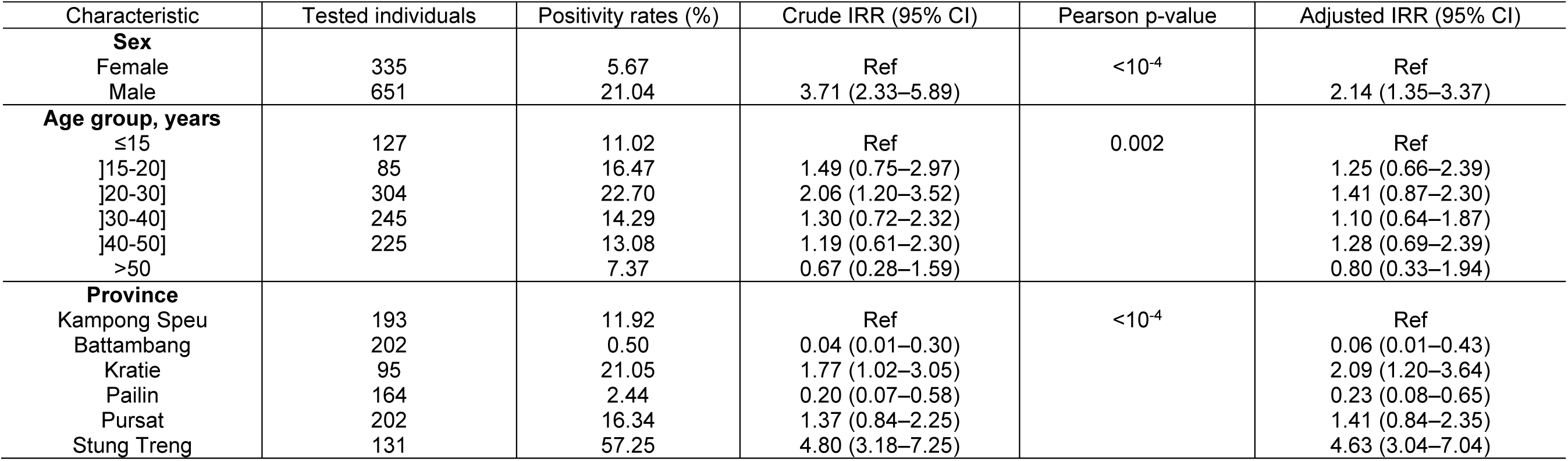
Crude and adjusted infection risk ratios (IRR) for malaria infection.

The proportion of microscopy positive samples among all qPCR positives was 78.8% (123/156, 95% CI: 71.8-84.5%), accordingly 21.2% were sub-microscopic infections. Among the qPCR positive participants not pre-diagnosed by VMW, 55.9% (38/68, 95% CI: 44.1-67.1%) were microscopy positive. Parasitemia among *Plasmodium* infected patients was not different between male and female patients (whether considering all patients or only those not pre-diagnosed by VMW) nor between the different age groups (considering all patients or only those not pre-diagnosed by VMW) (Table 2). Parasitemia among qPCR positive patients varied significantly across HCs. When considering all infected patients; median parasitemia differed significantly between HCs (Kruskal-Wallis, p=0.0006). Similar differences were observed when restricting the analysis to patients not pre-diagnosed by VMW (p=0.0370) (Table 3).

At the health-center level, qPCR positivity rates were positively associated with mean parasitemia measured among infected patients (Spearman, r=0.76, p=0.0414) (Figure 3). This association remained significant when restricting the analysis to patients not pre-diagnosed by VMW (Spearman, r=0.90, p=0.0054). Health centers with the lowest qPCR positivity rates (<3%, Boeung Run, Krachab and Aoral) exhibited significantly lower parasitemias among qPCR positive patients, compared to HCs with higher infection rates (>16%, Siem Pang, Kampong Cham, Kravanh and Trapaing Cho), both when considering all patients and when restricting to patients not pre-diagnosed (Mann Whitney, p<0.0001 and p=0.0085, respectively) (Figure 3.).

**Figure 3.**
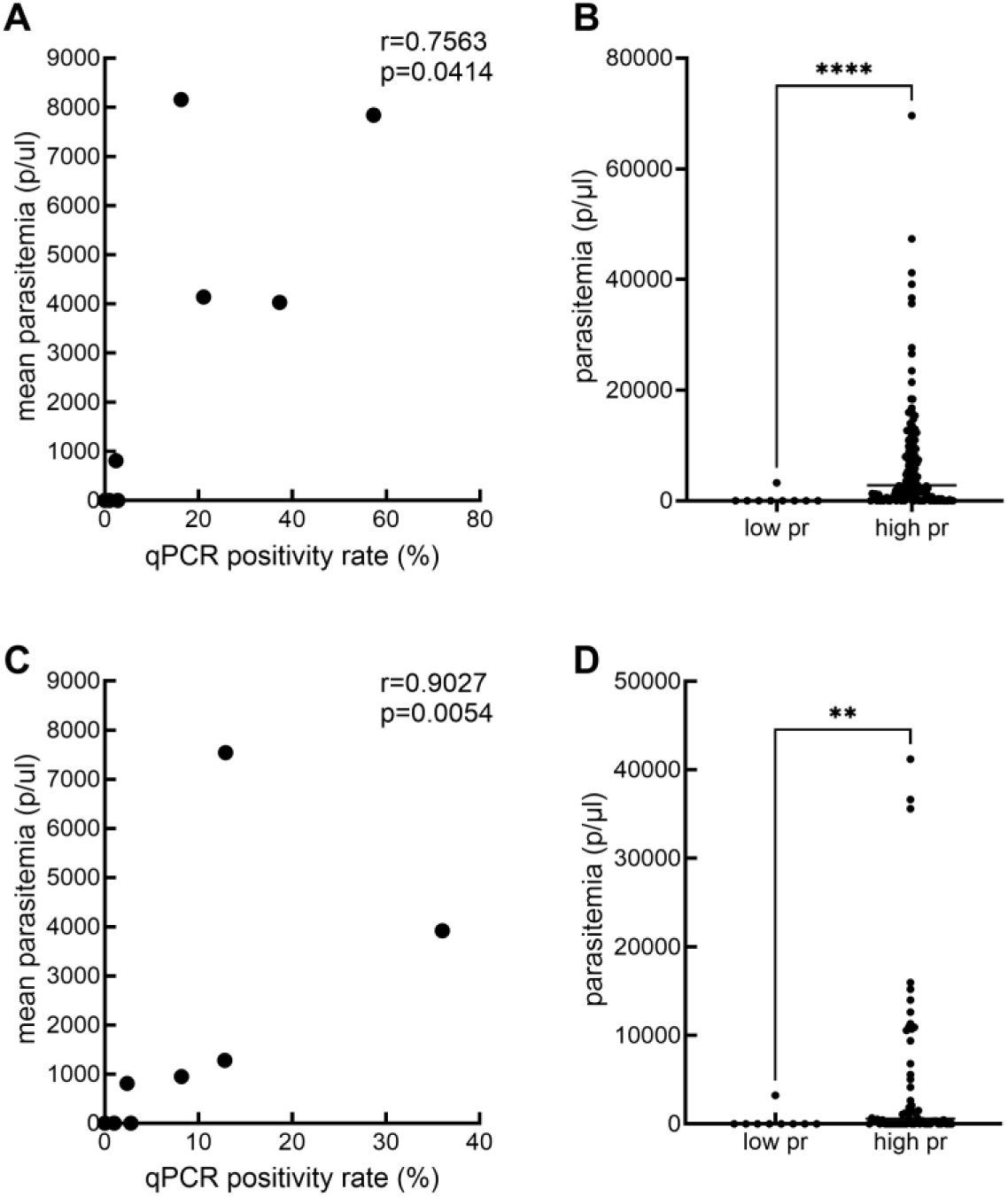
Panels (A) and (C): Correlation between mean parasitemia (parasite/μl) quantified by microscopy and qPCR positivity rates of *Plasmodium* infections in all infected patients (A) or in those not pre-diagnosed by VMW (C). Each dot represents one HC. Panels (B) and (D): Distribution of parasitemia in patients from low positivity rate HCs (Boeung Run, Krachab and Aoral) and from higher positivity rate ones (Siem Pang, Kampong Cham, Kravanh and Trapaing Cho) in all infected patients (B) or in those not pre-diagnosed by VMW (D). Each dot represents one patient. pr: qPCR positivity rate.

To account for small sample sizes at the health-center level and for left-censoring of parasite density measurements due to the detection limit of microscopy we conducted an additional sensitivity analysis using a Bayesian hierarchical censored model. Posterior median parasitemia estimated for each HC showed a consistent positive association with qPCR positivity rates, supporting the robustness of the observed pattern appropriately reflecting uncertainty (Supplementary Figure 2.).

*Plasmodium* species were successfully identified for 99% (154/156) of infections. All human malaria parasites were identified across the study sites(Figure 4). The majority of infections were caused by *P. vivax* diagnosed either alone (75%, 116/154) or in mixed infections with *P. malariae* (7.8%, 12/154). For two patients (1.3%, 2/154) a triple infection with *P. vivax*, *P. malariae* and *P. ovale* was detected. Single *P. malariae*, *P. knowlesi* or *P. falciparum* were identified in 5.8% (9/154), 5.8% (9/154) and 3.9% (6/154) of patients, respectively. *P. malariae* (either single or mixed infections) was detected in patients from western Cambodia (Pursat and Kampong Speu) as well as in patients from eastern Cambodia (Kratie). Similarly, *P. falciparum* was identified in patients from western Cambodia (Pailin, Pursat and Kampong Speu) and eastern Cambodia (Stung Treng). *P. knowlesi* was only identified in patients from western Cambodia (Pailin and Pursat) and the two *P. ovale* identified in mixed infections were both in patients from Kratie.

**Figure 4.**
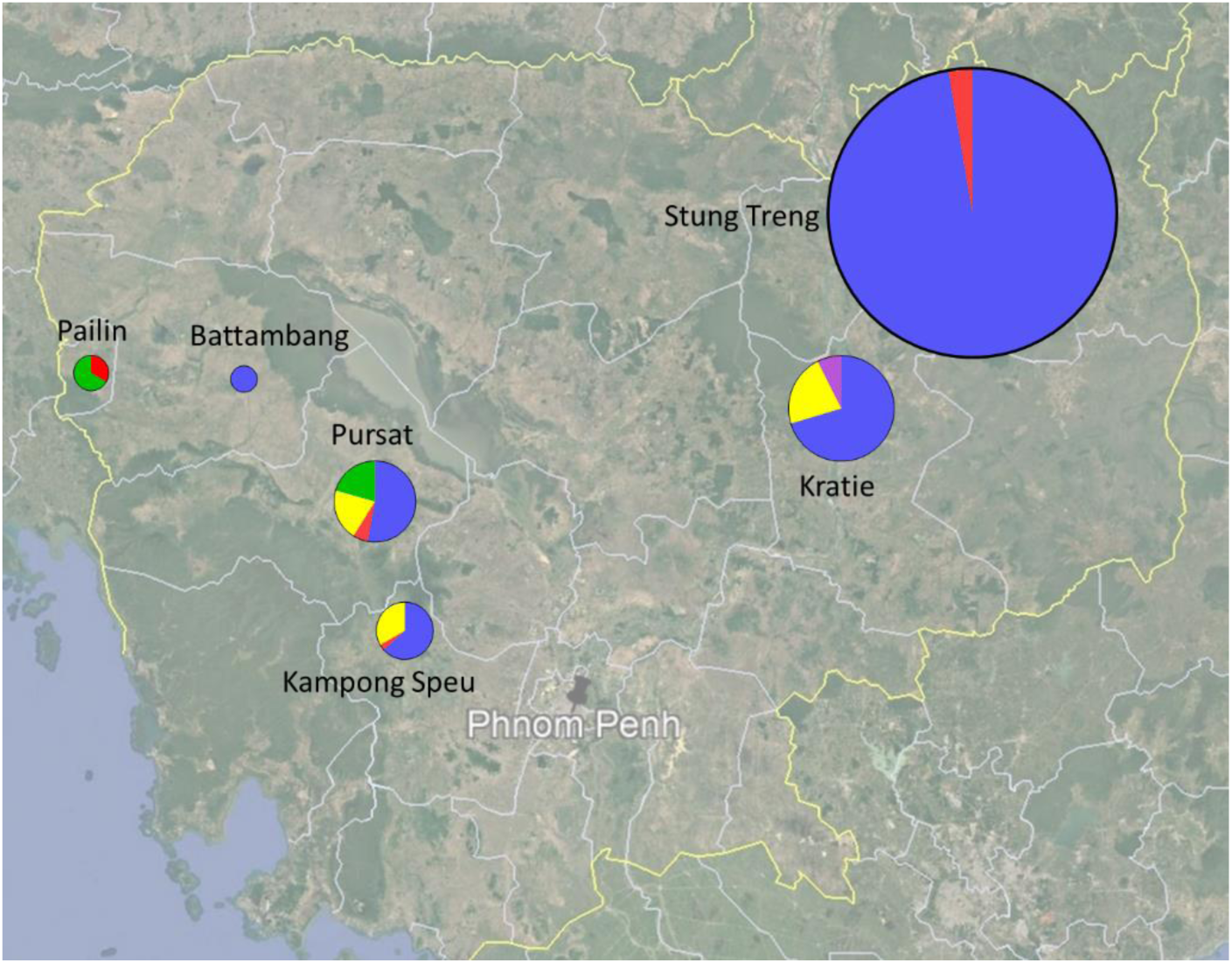
Distribution of the different *Plasmodium* species identified by qPCR across all HC. The size of the pie chart is proportional to the overall *Plasmodium* qPCR positivity rate measured in each province. Note that pie charts for Pailin and Battambang have been resized to be visible (3 and 10 times, respectively). Color 281 codes: blue *P. vivax*, red *P. falciparum*, green *P. knowlesi*, yellow *P. malariae*, purple *P. ovale*. Species of mixed infections are included separately.

To evaluate the performance of RDTs and microscopy to diagnose *Plasmodium* infections, we considered only patients not pre-diagnosed using RDTs by VMW. Overall, 898 patients, of which 68 (7.6%, 95% CI: 6.0-9.5%) were infected by *Plasmodium*, were included in the analysis (Table 5). Microscopy specificity could not be assessed as it was performed only on qPCR positive samples but its sensitivity was 55.9% (95% CI: 44.1-67.1%). The sensitivity of RDT to detect *Plasmodium* infections (any species) was 39.7% (95% CI: 28.9-51.6%) and its specificity was 100% (95% CI: 99.5-100%). Positive predictive value (PPV) was 100% (95% CI: 87.5-100%) while negative predictive value (NPV) was 95.3% (95% CI: 93.7-96.5%) and accuracy was 95.4% (95% CI: 94.1-96.8%). The proportion of RDT true positive samples was significantly different between *Plasmodium* species (Fisher, p=0.0011) with 40% (95% CI: 26-56%) of *P. vivax* infections, 44% (95% CI: 19-73%) of *P. malariae*, 86% (95% CI: 49-99%) of *P. knowlesi* and 75% (95% CI: 30-99%) of *P. falciparum* infections diagnosed by RDTs. None of the mixed *P. vivax*-*P. malariae* or *P. vivax*-*P. malariae*-*P. ovale* infections were detected by RDTs (0%, 95% CI: 0-26%).

**Table 5.**
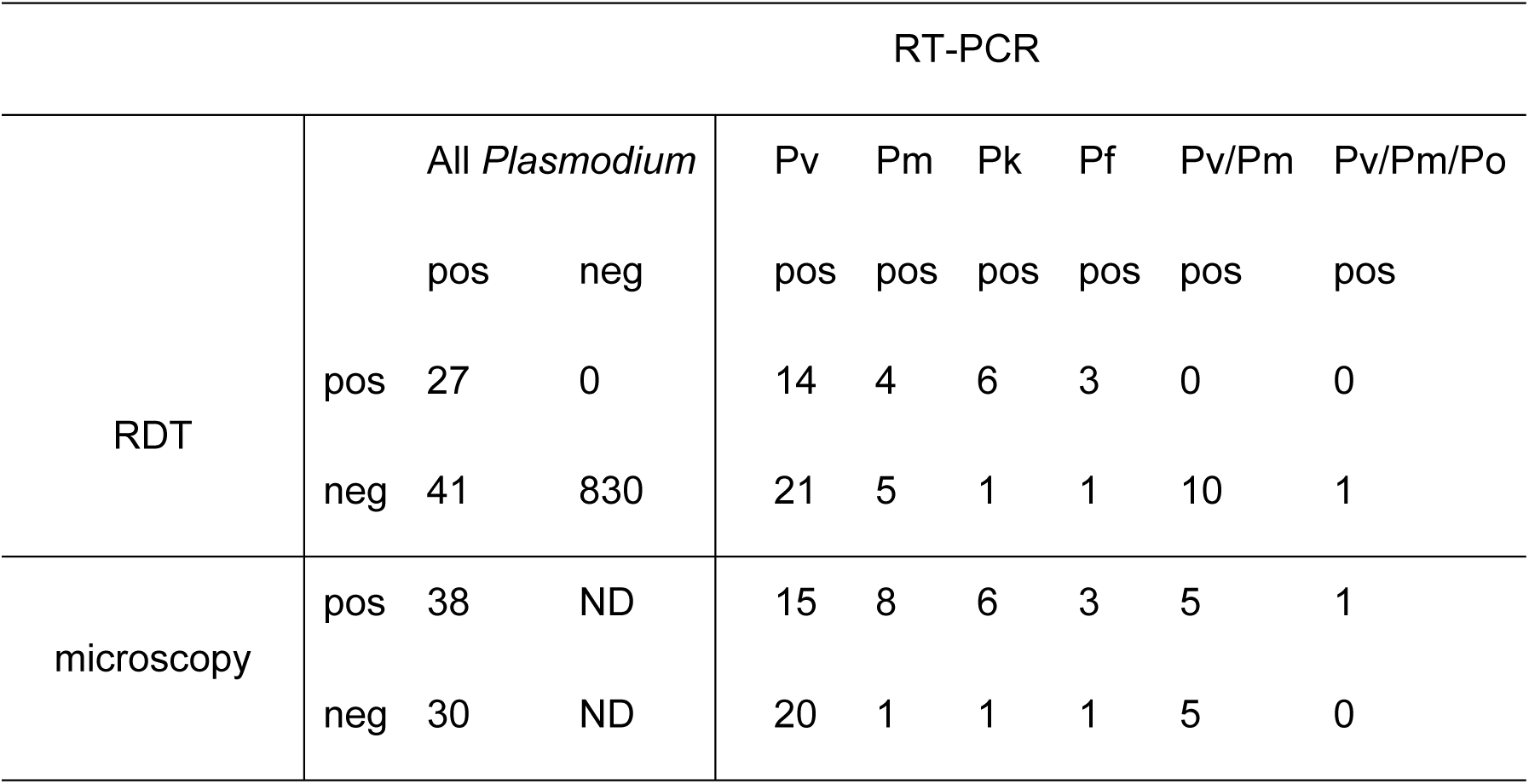
Performance of RDT and microscopy to diagnose *Plasmodium* infections compared to qPCR as gold standard for patients not pre-diagnosed by VMW.

Parasitemia in qPCR positive samples for all *Plasmodium* species considered was significantly lower in RDT negative samples (median 0 parasites/μl, IQR: 0-502, range: 0-10744) compared to RDT positive samples (5047 parasites/μl, IQR: 1105-12652, range: 0-41222, Mann-Whitney, p<0.0001) (Figure 5). This difference was also significant for infections caused by *P. vivax* (RDT negative median: 0 parasites/μl, IQR: 0-0, range: 0-2090, RDT positive median: 4597 parasites/μl, IQR: 1116-11085, range: 0-41222, p<0.0001). Although sample size was too small for statistical testing, a similar trend was observed for *P. falciparum* and *P. knowlesi* infections where, for both species, the only RDT negative infection was sub-microscopic. However, for *P. malariae* infections, there was no significant difference in parasitemia between RDT positive (median: 738 parasites/ul, IQR: 333-4459, range: 321-5577) and RDT negative (1293 parasites/μl, IQR: 268-10663, range: 0-10744, p=0.5556) infections. All 10 mixed *P. vivax*-*P. malariae* infections were RDT negative while half were submicroscopic and the *P. vivax*-*P. malariae*-*P. ovale* mixed infection was RDT negative and had a parasitemia of 468 parasites/ul.

**Figure 5.**
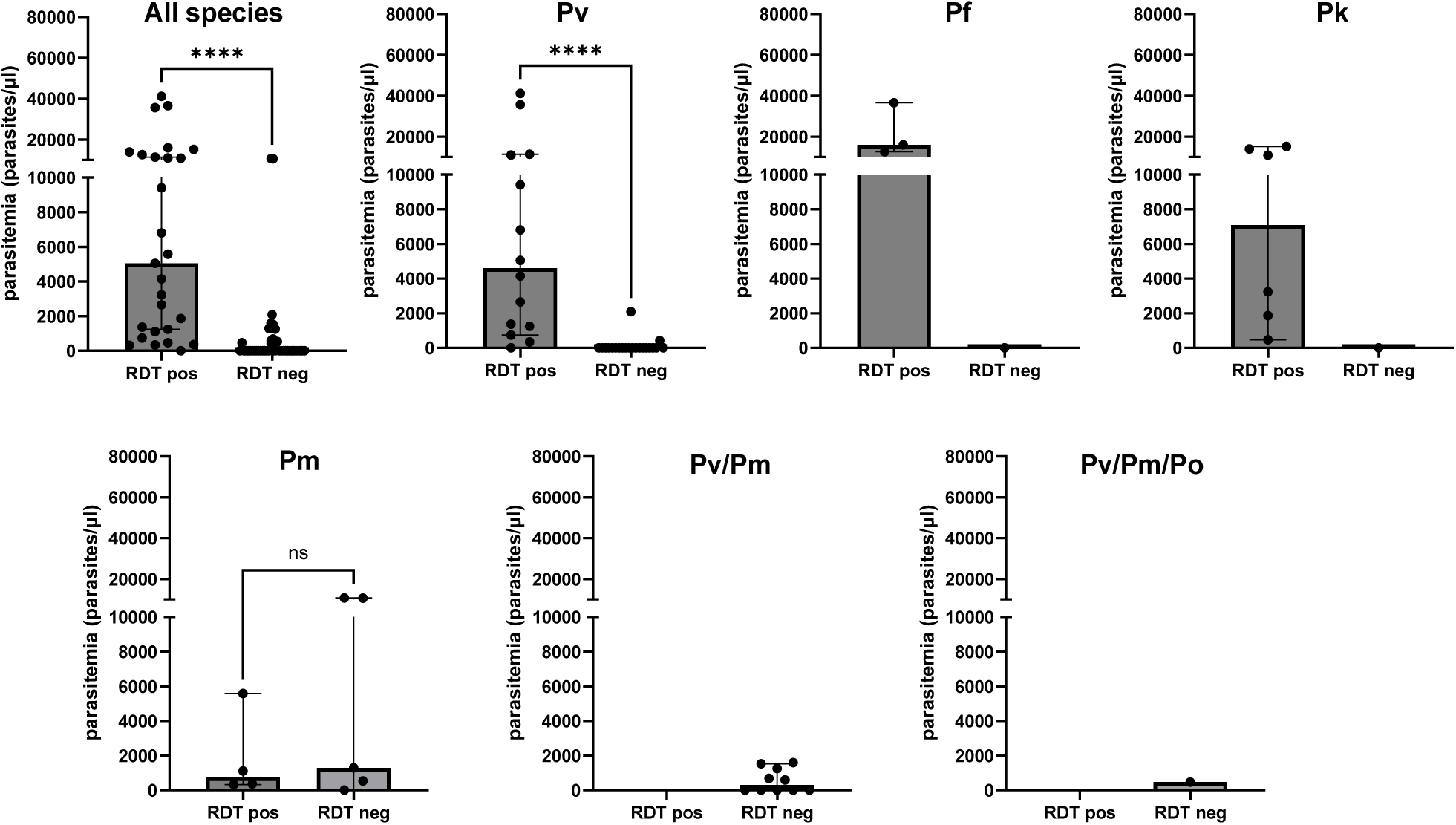
Parasitemia measured in *Plasmodium* infected samples diagnosed by qPCR and compared between RDT positive and negative samples. Pv: *P. vivax*, Pm: *P. malariae*, Pk: *P. knowlesi*, Pf: *P. falciparum*, Po: *P. ovale.* Each dot represents one patient.

Among infections not pre-diagnosed by VMW, there was a significant difference in the proportion of RDT true positive tests between the different provinces (Fisher, p=0.0060) and between the different HC (p=0.0118). The median true positive rates by RDTs in HC with the lowest qPCR positivity rates (<3%, Boeung Run, Krachab and Aoral) was 0% (95% CI: 0-25%) while median true positive rates in HC with higher qPCR positivity rates (>16%, Siem Pang, Kampong Cham, Kravanh and Trapaing Cho) was 34% (95% CI: 22-68%), although the difference did not reach significance (Mann Whitney, p=0.1143, Supplementary Figure 2).

## DISCUSSION

We describe here a cross-sectional survey conducted over an entire malaria transmission season in 8 health facilities located in 6 provinces of Cambodia with current or historical records of malaria endemicity. Our study shows that *Plasmodium* infections were detected using molecular diagnostics in febrile patients from all provinces. However, marked differences in positivity rates were observed between regions, with Stung Treng having the highest burden of malaria infections and Battambang the lowest one. These differences are in line with reported cases diagnosed at health facilities using RDTs and informed in the national malaria registry. We also confirm the risk factors known to be associated with malaria infections, namely being a young adult male, at higher risk due to occupational behavior (5,8,25,26).

Our work, however, reveals a huge proportion of infections among treatment-seeking individuals missed by current RDTs, which lead to clinical diagnostic failures and untreated carriers. Although the proportion of false negative RDTs varied significantly between the different species, the poor performance was particularly observed for the most prevalent *P. vivax* and *P. malariae* infections. In our work, all species known to infect humans were detected, including *P. knowlesi*. Reassuringly, as *P. knowlesi* is closely related to *P. vivax*, the majority of infections were picked up by RDTs (although identified as *P. vivax*) and patients received antimalarial treatment, highlighting the utility of a *P. vivax* LDH component in RDTs to detect this species. The only false negative RDT test for *P. knowlesi* was a submicroscopic infection, a finding in line with a recent study evaluating RDTs to diagnose simian Plasmodium infections (27). However, for *P. malariae*, and surprisingly for all mixed infections (whether *P. vivax*-*P. malariae* or *P. vivax*-*P. malariae*-*P. ovale*), RDTs performed particularly poorly with only 4 out of 20 infections testing positive. Perhaps RDTs with a pan-species LDH could be of interest to improve this performance, although this should be evaluated notably against these neglected malaria species. Regarding *P. falciparum*, our molecular diagnostic confirms current trends reported in Cambodia with near elimination of *P. falciparum* as only 6 cases were detected across the whole study period and sites.

In a previous study conducted in the Brazilian Amazon among predominantly asymptomatic individuals, a positive association between *Plasmodium* prevalence and parasite densities was observed (28). We report a similar pattern among symptomatic patients in the present study. In Cambodia, in a near-elimination setting, we observed that parasitemia among symptomatic, treatment-seeking patients tended to be lower in health centers with very low *Plasmodium* positivity rates compared to centers with higher positivity rates. This pattern was consistently observed across analytical approaches and remained apparent when excluding patients pre-diagnosed at the community level.

Although this study was not designed to establish causal relationships at the individual level, it aimed to explore whether consistent epidemiological patterns could be identified across HCs with contrasted transmission intensity. Health centers represent a relevant operational unit in the Cambodian malaria control program, as transmission intensity, diagnostic practices, and patient catchment areas are structured at this level.

Although it is not investigated within the scope of this study, a possible mechanism for this observation could be that in low transmission areas undergoing significant malaria elimination efforts, only individuals with significant immunity are still infected by parasites, therefore reducing parasitemia. Alternatively, it could also be hypothesized that in areas with intense malaria control efforts leading to near elimination, only parasites with intrinsic lower virulence causing low parasitemia infections remain. Further studies looking notably at parasite genetic population structure and host immunology could help determine if this is a valid hypothesis.

This analysis has several limitations. The number of health centers included was limited, and the number of qPCR-positive samples was modest. Parasite densities were measured only among qPCR-positive individuals. Moreover, parasitemia measurements were subject to left-censoring due to the detection limit of microscopy. To address these constraints, we conducted sensitivity analyses using a Bayesian hierarchical censored model, which supported the robustness of the observed pattern while appropriately reflecting uncertainty.

Despite these limitations, the consistency of findings across analytical approaches and their biological plausibility suggests that declining parasite densities in low-transmission settings may represent a challenge for malaria diagnosis. Furthermore, our findings, derived from predominantly non-*falciparum* infections, align with similar observations reported for *P. falciparum* cases (29).

## CONCLUSION

Altogether, these results show that upon significant reduction of malaria from the country, in areas with very low number of reported cases, misdiagnosis and false negative RDTs are likely to increase in treatment-seeking individuals as elimination progresses. Molecular diagnostics of infections for patient care in these remote rural areas are likely impractical. However, blood collection from RDT-negative individuals presenting at HCs, and retrospective batch molecular testing should be considered to be established as part of the national strategy to achieve elimination. Such surveillance would also have the benefit to contribute to the prevention of re-establishment of malaria once eliminated from an area.

Overall, this work highlights the challenges of diagnosing *Plasmodium* infections in febrile individuals using RDTs (and microscopy) in the context of near elimination and warrants for improved diagnostic sensitivity.

## Supporting information

Supplementary_material_Khim_et_al

## Data Availability

All data produced in the present study are available upon reasonable request to the authors.

## DECLARATIONS

### Availability of data and materials

The datasets used and/or analysed during the current study are available from the corresponding author on reasonable request.

### Authors’ contributions

JP designed the study. NKh, AO, ST, SS, SG, LBFD, VE, ME, SC, CS, RE, CKh, CKu, NKl, SK were responsible for data collection. DL and JP oversaw the study. NKh, AO, CF, MW and JP did the data analysis. NKh, AO and JP wrote the first draft.

## Acknowledgments

We are grateful to the health facility staff of participating HC as well as to the patients who accepted to participate in this study. This study was funded by Institut Pasteur du Cambodge core funding. JP is supported by the NIH/NIAID (R01AI173171, R01AI175134 and R61AI187100). MW and JP are supported by the Pasteur International Unit PvESMEE. Funders had no role in the study’s design, data collection, analysis, interpretation, or report writing.

