## Supplementary_material_Khim_et_al for "Challenges in Plasmodium diagnostics in countries nearing malaria elimination: a cross-sectional survey among treatment-seeking patients in health facilities in malaria endemic provinces of Cambodia with contrasted transmission intensity"

**Supplementary Table 1.** Primer sequences and annealing temperatures of *Plasmodium* species specific PCR assays

| ***Plasmodium* gene target** | **Primer name** | **Sequence (5’-3’)** | **Used for** | **Expected product size** | **Annealing temp.** | **Reference** |
| --- | --- | --- | --- | --- | --- | --- |
| *Plasmodium* *cytochrome*  *b* gene | RTPCRScreening2_F | TGGAGTGGATGGTGTTTTAGA | Real-time PCR screening | 211 bp | 60^0^C | Canier et al., Malar J. 2013 |
|  | RTPCRScreening2_R | TTGCACCCCAATARCTCATTT |  |  |  |  |
|  | RTPCRScreening2_F | TGGAGTGGATGGTGTTTTAGA | Primary PCR species/ Sanger sequencing | 400 bp | 58^0^C |  |
|  | RTPCRSreening3_R | ACCCTAAAGGATTTGTGCTACC |  |  |  |  |
|  | Pf_RTPCR_F | ATGGATATCTGGATTGATTTTATTTATGA | Nested real-time  PCR Pf | 154 bp | 62^0^C |  |
|  | Pf_RTPCR_R | TCCTCCACATATCCAAATTACTGC |  |  |  |  |
|  | Pv_RTPCR_F | TGCTACAGGTGCATCTCTTGTATTC | Nested real-time  PCR Pv | 165 bp | 62^0^C |  |
|  | Pv_RTPCR_R | ATTTGTCCCCAAGGTAAAACG |  |  |  |  |
|  | Pm_RTPCR_F | ACAGGTGCATCACTTGTATTTTTTC | Nested real-time  PCR Pm | 213 bp | 62^0^C |  |
|  | Pm_RTPCR_R | TGCTGGAATTGAAGATAATAAATTAGTAATAACT |  |  |  |  |
|  | Po_RTPCR_F | GTTATATGGTTATGTGGAGGATATACTGTT | Nested real-time  PCR Po | 82 bp | 62^0^C |  |
|  | Po_RTPCR_R | CGAATGGAAGAATAAAATGTAGTACG |  |  |  |  |
|  | CYTB_PRIM_1F | AACAGGTGTATTTTTAGCAAGTCG | Primary PCR Pk/ Sanger sequencing Pk | 482 bp | 58^0^C | This study |
|  | CYTB_PRIM_3R | ACCCTAAAGGATTTGTGCTACC |  |  |  |  |
|  | Pk_RTPCR_MAR_F | CTCCAGAAATTTCTTACGCATACTAC | Nested real-time  PCR Pk | 240 bp | 64^0^C |  |
|  | Pk_RTPCR_MAR_R | CCCAAGGTAAAACATAACCTATAAAA |  |  |  |  |
|  | CYTB_PCR_F | TGTAATGCCTAGACGTATTCC | Primary PCR | 1200 bp | 53^0^C | Muehlenbein et al., Molecular  biology and evolution. 2015 |
|  | CYTB_PCR_R | GTCAAWCAAACATGAATATAGAC |  |  |  |  |
|  | Cytb_nested_F | TCTATTAATTTAGYWAAAGCAC | Nested PCR Pk/ Sanger sequencing Pk | 1109 bp | 61^0^C |  |
|  | Cytb_nested_R | GCTTGGGAGCTGTAATCATAAT |  |  |  |  |
| *18s rRNA* gene | rPLU1 | TCAAAGATTAAGCCATGCAAGTGA | Primary PCR | 1639 bp | 58^0^C | Snounou et al., Methods Mol Med. 2002 |
|  | rPLU5 | CCTGTTGTTGCCTTAAACTCC |  |  |  |  |
|  | rUNF1 | TTAAGCCATGCAAGTGAAAGTAT | Nested PCR Pk/ Sanger sequencing Pk | 1034 bp | 55^0^C | Dixit et al., PLOS Neglected Tropical Diseases. 2018 |
|  | rUNR1 | CGGTATCTGATCGTCTTC |  |  |  |  |
| The details of RT-PCR assays (positive/negative classification criteria, controls, number of repeats, etc.) are described in details in Canier et al., Malar J. 2013 (doi: 10.1186/1475-2875-12-405). The assessors of RT-PCR assays were blinded to the RDT results. | | | | | | |

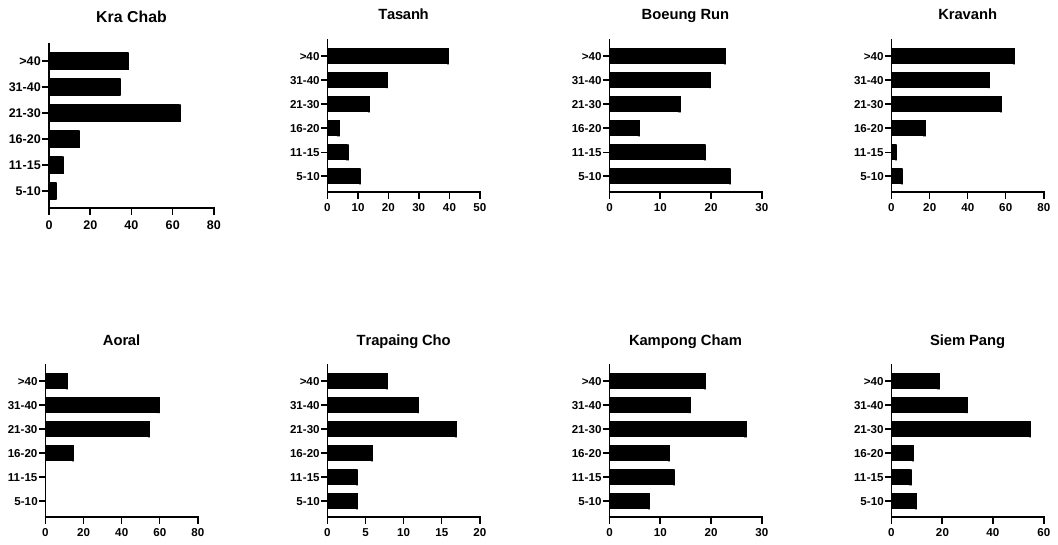

**Supplementary Figure 1**. Age distribution of patients attending participating HCs and enrolled in this study.

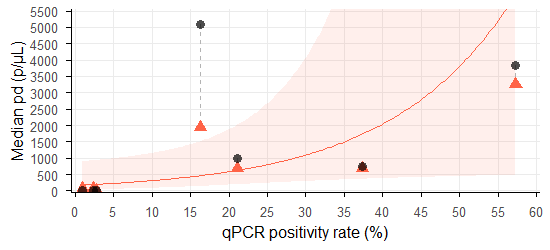

A

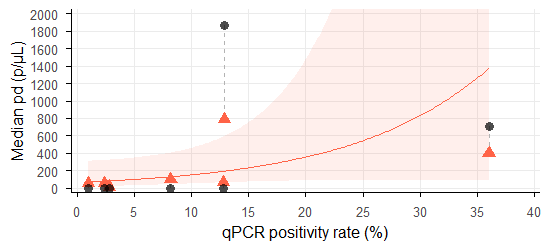

B

**Supplementary Figure 2.** HC-level median parasite densities were estimated using a Bayesian hierarchical censored model, to account for the limit of detection of microscopy and small sample sizes. The association between posterior medians and qPCR positivity rate was assessed via Spearman correlation. In the case of all qPCR positive samples Spearman correlation between posterior median parasite density and qPCR positivity rate was 0.75 (95% credible interval: 0.25 to 0.89), with a 99.6% posterior probability that the correlation is positive, indicating a moderate-to-strong association (Panel A). In case of samples non pre-diagnosed the correlation was slightly weaker 0.61 (95% credible interval: -0.11 to 0.93), with a 94.8% posterior probability that the correlation is positive, due to the more limited sample size (Panel B). A GLM regression line was fitted for visualization, showing the relationship and 95% confidence interval, but no inference was drawn from this regression. Red triangles indicate the posterior median parasite densities per HC, black dots indicate the raw median parasite densities.

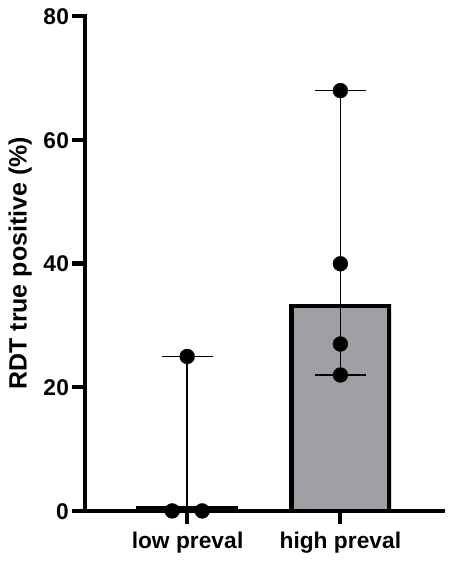

**Supplementary Figure 3.** RDT true positive rates among qPCR diagnosed *Plasmodium* infections in patients not-pre-diagnosed by VMW in the low prevalence HCs (Boeung Run, Krachab and Aoral) and in high prevalence HCs (Siem Pang, Kampong Cham, Kravanh and Trapaing Cho).
